# Contraception in breast cancer survivors from the FEERIC case-control study (performed on behalf of the Seintinelles research network)

**DOI:** 10.1101/2021.12.15.21267867

**Authors:** Clara Sebbag, Christine Rousset-Jablonski, Florence Coussy, Isabelle Ray-Coquard, Clémentine Garin, Clémence Evrevin, Aullène Toussaint, Marion Cessot, Julie Labrosse, Lucie Laot, Lauren Darrigues, Angélique Bobrie, William Jacot, Claire Sénéchal-Davin, Marc Espié, Sylvie Giacchetti, Patrick Charveriat, Geneviève Plu-Bureau, Lorraine Maitrot-Mantelet, Anne Gompel, Charles Chapron, Pietro Santulli, Bernard Asselain, Judicaël Hotton, Charles Coutant, Julien Guerin, Christine Decanter, Audrey Mailliez, Lidia Delrieu, Elise Dumas, Laura Sablone, Research Network Seintinelles, Fabien Reyal, Anne-Sophie Hamy

**Affiliations:** Department of Medical Oncology, Institut Curie, Paris, France; University Paris, Paris France; Residual Tumor & Response to Treatment Laboratory, RT2Lab, Translational Research Department, INSERM, U932 Immunity and Cancer, University Paris, Paris, France; INSERM U1290 RESearch on HealthcAre PErformance (RESHAPE), University Claude Bernard Lyon 1, Lyon, France; Department of Surgical Oncology, Centre Léon Bérard, Lyon, France; Department of Surgical Oncology, Institut Curie, Paris, France; University Paris, Paris France; Department of Medical Oncology, Institut du cancer de Montpellier, Montpellier, France; Department of Oncogenetics, Institut Bergonié, Bordeaux, France; Sénolopole, Hôpital Saint Louis, AP-HP, Paris, France; University Paris, Paris France; Department of Gynecology, Hôpital Cochin, Paris, France; University Paris, Paris France; Department of Biostatistics, Unicancer, Paris, France; Department of Surgical Oncology, Institut Godinot, Reims, France; Department of Surgical Oncology, Centre Georges-François Leclerc - Unicancer, Dijon, France; Clinical Research Department, Centre Georges-François Leclerc - Unicancer, Dijon, France; Data Factory, Data Office, Institut Curie, 25 rue d’Ulm, 75005 Paris, France; ART department, Centre Hospitalier Regional Lille, Lille, France; Department of Medical Oncology, Centre Oscar Lambert, Lille, France; Seintinelles Research Network, 40 Rue Rémy Dumoncel, 75014 Paris

**Keywords:** contraception, breast cancer, survivorship, contraceptive counseling, emergency contraception

## Abstract

**Objective:** To compare the prevalence of contraception in breast cancer (BC) patients at risk of unintentional pregnancy (*i*.*e*. not currently pregnant or trying to get pregnant) and matched controls.

**Design:** The FEERIC study (Fertility, Pregnancy, Contraception after BC in France) is a prospective, multicenter case-control study. Data were collected through online questionnaires completed on the Seintinelles* research platform.

**Setting:** Not applicable

**Patient(s):** BC patients aged 18-43 years, matched for age and parity to cancer-free volunteer controls in a 1:2 ratio.

**Intervention(s):** None

**Results:** In a population of 1278 women at risk of unintentional pregnancy, the prevalence of contraception at study inclusion did not differ significantly between cases (340/431, 78.9%) and controls (666/847, 78.6%, *p*=0.97). However, the contraceptive methods used were significantly different, with a higher proportion of copper IUD use in BC survivors (59.5% *versus* 25.0% in controls *p*<0.001). For patients at risk of unintentional pregnancy, receiving information about chemotherapy-induced ovary damage at BC diagnosis (OR= 2.47 95%CI [1.39 - 4.37] and anti-*HER2* treatment (OR=2.46, 95% CI [1.14 - 6.16]) were significantly associated with the use of a contraception in multivariate analysis.

**Discussion:** In this large French study, BC survivors had a prevalence of contraception use similar to that for matched controls, though almost one in five women at risk of unintentional pregnancy did not use contraception. Dedicated consultations at cancer care centers could further improve access to information and contraception counseling.

## Introduction

Breast cancer (BC) is the most common cancer in women. Approximately 11000 women under the age of 45 years are diagnosed with BC annually in France (Defossez et al., 2019). Survival is increasing, and more attention is now being paid to the adverse effects of treatment. In particular, over the last decade, physicians have begun to pay more attention to the possibility of pregnancy following BC. Fertility preservation procedures are gradually being incorporated into routine care, together with prophylactic treatments to prevent damage to the ovaries during chemotherapy.

Concerns have been raised over decades about the impact on recurrence of breast cancer after a pregnancy and BC patients have long been advised against conception in the future, due to fears that pregnancy could adversely affect their breast cancer outcome. However, many data have since emerged to indicate that pregnancy does not have a detrimental effect on survival (Azim et al., 2011), regardless of ER status (Azim et al., 2013; Lambertini et al., 2018), and the presence or absence of a germline *BRCA* mutation (Lambertini et al., 2020).

Contraception after BC has been little studied. However, it is a particularly important issue because pregnancy planning in these patients is crucial from a medical point of view, as highlighted in a previous study (Han et al., 2015). Patients who do not wish to become pregnant should actively avoid pregnancy, particularly during tamoxifen treatment, as this drug is known to have potential teratogenic effects (Barthelmes and Gateley, 2004). Moreover, chemotherapy-induced amenorrhea might be associated with an unpredictable resumption of menses, potentially resulting in an unwanted pregnancy. Effective, safe, well-tolerated contraception is therefore of considerable importance for this population.

Female hormonal contraception has been available for over 50 years and is used by more than 300 million women worldwide(United Nations, 2019). Hormonal contraceptive (combined estroprogestin or progestin pill, progestin implant, vaginal ring, transdermal patch, and levonorgestrel intrauterine system) are classically contraindicated in BC patients, and women are generally advised to stop hormonal contraceptive use at the time of BC diagnosis. Classic options for alternative contraceptive methods, according to guidelines (WHO, 2015), include intrauterine device (IUD), or barrier methods. However, IUD use may be limited by an inability to tolerate the device and abnormal bleeding patterns. It may also aggravate bleeding disorders during menstrual recovery from chemotherapy-induced amenorrhea. Thus, breast cancer survivors have very few contraceptive options, mostly non-hormonal methods.

Several studies have demonstrated a positive impact of contraceptive counseling on women’s acceptance of contraception (Gaudet et al., 2004), but few data have been published concerning the effect of contraceptive counseling in women with cancer. Previous studies have shown that sexually active cancer survivors have lower rates of use of World Health Organization tier I-II contraceptive methods (Dominick et al., 2015; Güth et al., 2015; Maslow et al., 2014), and are considered at high risk of unintended pregnancy (Quinn et al., 2014). To our knowledge, few studies have investigated the knowledge and use of emergency contraception in BC patients. In the FIRST cohort (Dominick et al., 2015), breast cancer patients were found to be three times less likely to use emergency contraception than other cancer survivors. The use of family planning services increased the use of tier I–II contraceptive methods by a factor of two and the use of emergency contraception by a factor of five (Dominick et al., 2015) to six (Maslow et al., 2014).

The FEERIC study was launched in France in March 2018 and was designed to compare fertility, pregnancy and contraception outcomes in young BC survivors and matched cancer-free women. The objective of the study described here was to analyze contraception use at BC diagnosis and during follow-up, and to compare contraceptive use between BC survivors and matched controls.

## Materials and methods

### Study design and data collection

The design of The FEERIC (Fertility, Pregnancy, Contraception after BC in France) study has been described elsewhere (Mangiardi-Veltin, n.d.). Briefly, the FEERIC study is a prospective case-control study assessing the impact of BC treatment on fertility, pregnancy, and contraception in young BC survivors. Women were recruited from March 13, 2018 to June 27, 2019. Data were collected via self-administered online questionnaires released through the Seintinelles* research platform. Seintinelles* is a collaborative social network created in 2012 to accelerate the recruitment of French volunteers for cancer research studies, by connecting researchers with men and women of various ages, social and medical backgrounds, with or without a history of BC, willing to participate in research studies.

The scientific board of the Seintinelles approved the FEERIC project in December 2015, and the ethics board of Sud Ouest Outre Mer II approved the project on October 5, 2017.

### Study population

The inclusion criteria for cases were: female patients aged from 18 to 43 years with a previous diagnosis of localized, relapse-free BC (invasive or *in situ*) and who had completed treatment (surgery, chemotherapy, and radiotherapy) at the time of enrollment. The exclusion criteria were previous hysterectomy and/or bilateral oophorectomy and/or bilateral salpingectomy. The controls were women aged from 18 to 43 years, free from BC or other cancers, who had not undergone hysterectomy, bilateral oophorectomy or bilateral salpingectomy (Fig.S1).

We initially planned to match each BC patient (case) for age (± 2 years) and parity with two volunteers (controls) recruited prospectively within the Seintinelles network and one control from the patient’s close circle of friends and relatives. However, too few controls of this second type were recruited. We therefore pooled these controls with the volunteers recruited through the Seintinelles network, and each case was matched to two controls based on age and previous parity.

We excluded the women who were attempting to conceive or pregnant at inclusion (cohort 1 on the study flow chart), to define a subpopulation of women at risk of unintentional pregnancy (cohort 2).

### Contraception

#### Prevalence of contraception

The prevalence of contraception was defined as the percentage of women currently using, or whose sexual partner was currently using, at least one method of contraception, regardless of the method used.

#### Classification of contraceptive methods

Each contraceptive method was classified according to three classifications:

i. **The WHO contraceptive effectiveness tier classification** (Stanback et al., 2015) (World Health Organization et al., 2018); *Tier 1* consists of long-acting reversible contraception (LARC) (copper IUD, hormone containing intrauterine systems (hormonal IUS), implant) and non-reversible contraception (male and female sterilization). *Tier 2* consists of hormonal methods other than LNG-IUS (oral contraception, injection, patch, vaginal ring). *Tier 3* consists of barrier methods (male condom, diaphragm, cervical cap), and natural methods. Natural methods consist of the basal body temperature method, the cervical mucus method and the calendar method. *Tier 4* consists of female condoms, spermicides, and withdrawal.
ii. The **hormonal nature** of the contraception method (hormonal *versus* non-hormonal).

*Hormonal contraception* includes progestin-only (progestin-only mini pill, high-dose antigonadotrophic progestin, injectable progestins, implant, LNG-IUS) or combined hormonal contraception (combined oral contraceptive (COC), ring or patch). Conversely, the copper IUD, male or female condoms, natural methods, and other barriers methods were considered to be *non-hormonal contraceptive methods*. Women receiving gonadotropin-releasing hormone (GnRH) agonists in the context of their endocrine therapy were considered to be on contraception given the high gonadotropic suppressive potential of this treatment, but this method was not considered to be a hormonal contraceptive method as GnRH agonists do not formally deliver hormonal molecules.

(iii) The **reversibility** of the contraception method (definitive *versus* reversible): Essure*, tubal ligation and vasectomy were considered definitive, whereas all remaining methods were considered reversible.

For the sake of clarity, given the large number of contraceptive methods available, contraceptive methods are classified into three families (hormonal/non-hormonal/definitive), shown in Fig. 2B and Fig. 3A, and into five main classes (hormonal / copper IUD / male condom / definitive / others) in Fig. 2C.

### Emergency contraception

Emergency contraception was defined as methods of contraception (oral or IUD) used to prevent pregnancy after sexual intercourse. These contraceptive methods were not included in the previous classification of contraceptive methods, and data were collected in a dedicated section of the form.

### Study endpoint

The primary outcome measure was a comparison of the prevalence of contraception at inclusion between cases and controls, for women at risk of unintentional pregnancy.

### Statistical analysis

The study population is described in terms of frequencies for qualitative variables, or medians and associated ranges for quantitative variables. For analyses of the association between clinical variables (age, BMI, profession, study level, marital status, pregnancy history, comorbidity, comedications, family history of BC, gynecological follow-up, and theoretical need for contraception), prevalence of contraception, contraceptive counseling, emergency contraception and case-control status, we performed Student’s *t*-tests for continuous variables or Wilcoxon-Mann-Whitney tests for groups containing fewer than 30 patients, and for variables displaying multimodal distributions. Associations between categorical variables were assessed with Chi-squared tests, or with Fisher’s exact test if at least one category included fewer than three patients. A value of *p* ≤ 0.05 was considered statistically significant.

For identification of the factors predictive of contraceptive use in BC patients at risk unintentional pregnancy, variables were introduced into a univariate logistic regression model. A multivariate logistic model was then generated with a forward stepwise selection procedure, with all covariates included having a likelihood ratio test *p*-value ≤ 0.05. Data were processed and statistical analyses were performed with R software version 3.1.2 (www.cran.r-project.org, (R Foundation for Statistical Computing, 2009)).

## Results

### Contraceptive history and counseling at the time of BC diagnosis

At the time of BC diagnosis, 17.9% of the 517 patients (*n*=26) were pregnant or trying to conceive (*n*=67) and used no contraception. Of the remaining 424 patients, 383 (90.3%) were using contraception at BC diagnosis and a small subset of patients with no desire to have a child at all (9%) or no immediate desire to get pregnant (10%) did not use any contraception (Fig. 1A). For the 383 patients with contraception at diagnosis, 436 contraceptive methods were reported: two thirds of the methods used were hormonal (268/436, 61.5%), with combined oral contraceptives (*n*=131, 30%) and hormonal IUS the top two subtypes of hormonal contraception used (*n*=63, 14%); the remaining third of the methods were non-hormonal (168/436, 38.5%), with male condoms (*n*=92, 21%), copper IUD (*n*=42, 10%), and natural methods (*n*=29, 7%) as the top three methods (Fig. 1B).

**Figure 1:**
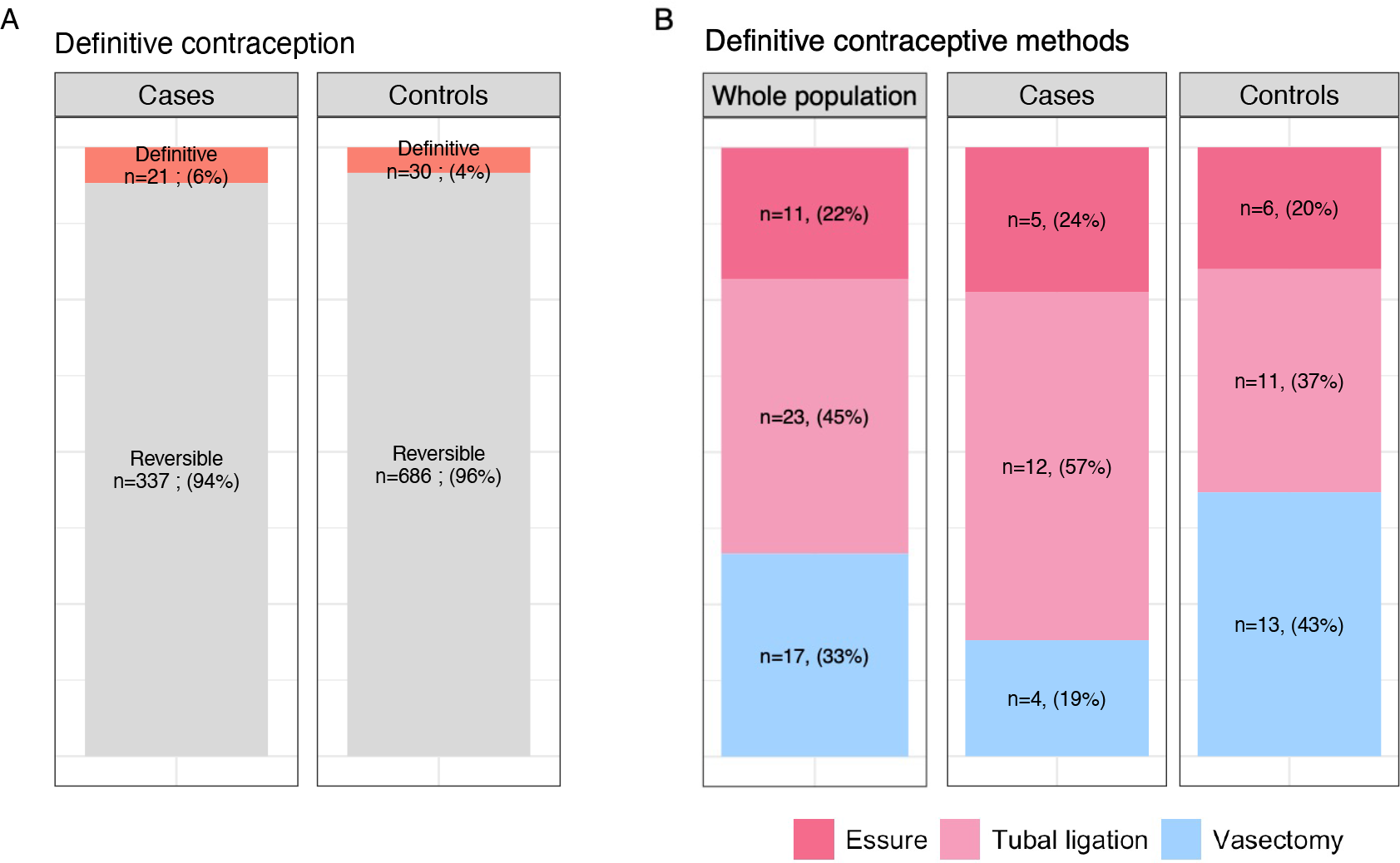
Contraceptive history and counselling at BC diagnosis. A, Contraception use according to attitudes to pregnancy at BC diagnosis. Data are reported per patient, and percentages are displayed for each category of attitude pregnancy. B, Contraception type at BC diagnosis (hormonal or non-hormonal contraception). Data are reported per contraceptive method (one patient can use several methods). C, Providers of contraception counseling at BC diagnosis. D, Rate of contraception discontinuation advice at BC diagnosis, for hormonal and non-hormonal contraception. Data are reported per contraceptive method (one patient can use several methods). Abbreviations: breast cancer (BC); combined oral contraceptive (COC); intrauterine system (IUS); intrauterine device (IUD); general practitioner (GP)

At BC diagnosis, specific contraceptive counseling was provided by a healthcare professional for 343 patients (66.3%). This counseling was mostly provided by oncologists (*n*=186, 54.2%) and gynecologists (*n*=80, 23.3%) (Fig. 1C). There was a high level of satisfaction with this information and the various contraceptive methods available (median score 9/10 and 8/10, respectively). Of 383 patients using contraception at diagnosis, 252 (65.8%) were asked to stop using at least one of their current methods of contraception. The proportion of the advice to discontinue differed between types of contraception (Fig. 1D), and was particularly high for women using oral hormonal contraception (248/265; 93.6%), including particularly, women using LNG-IUS (57/64; 89%).

### Changes between BC diagnosis and study inclusion

#### Changes in attitudes to the desire to become pregnant and attempts to conceive

Between BC diagnosis and study inclusion (median time: 34.9 months), 75.4% of the patients (390/517) had changed their attitude to pregnancy and/or their desire to become pregnant (Fig. 2A). In particular, 17 of the 218 patients (7.8%) with no previous desire to have a child at the time of diagnosis had changed their mindset and were trying to conceive. At study inclusion, the vast majority of patients were not attempting to conceive (*n*=431, 83.4%).

**Figure 2:**
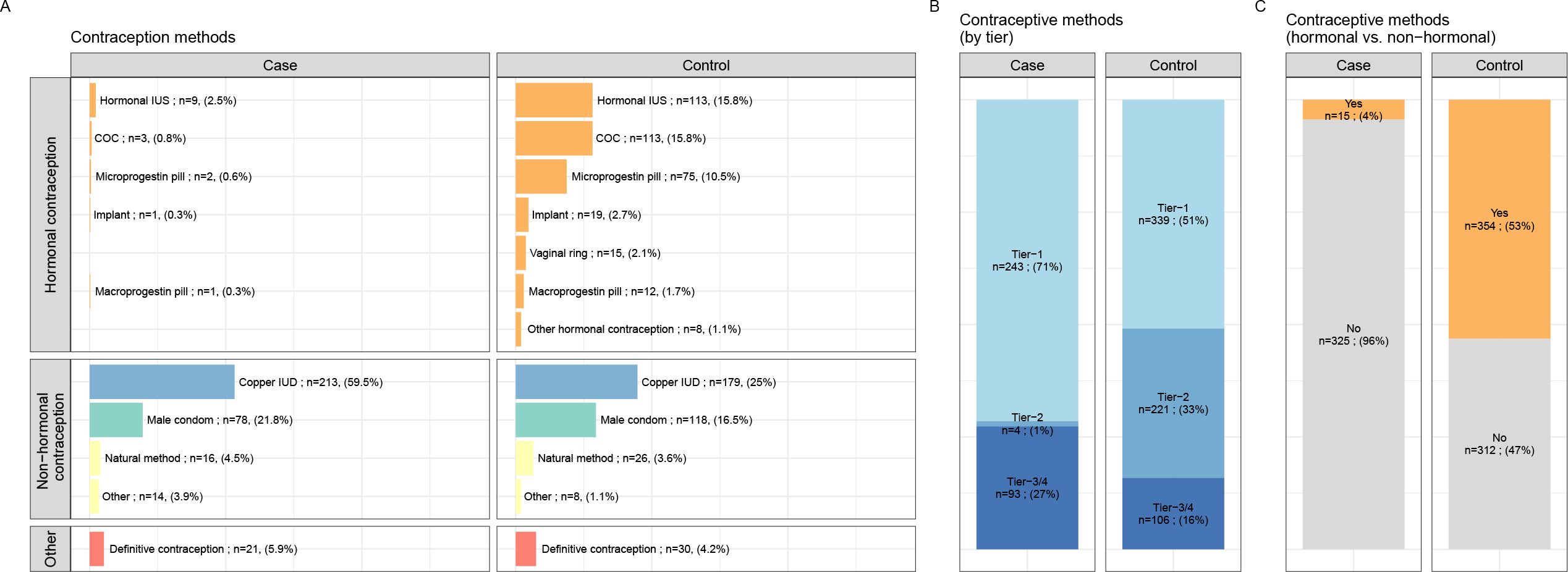
Changes from BC diagnosis to study inclusion in attitudes to pregnancy and contraceptive use A, Sankey plot of changes in attitude to and desire for pregnancy from diagnosis to study inclusion. Data are reported per patient. B, Sankey plot of changes in contraception methods, by class, from diagnosis to study inclusion. For the sake of clarity and readability, all hormonal contraceptives are grouped together, all definitive contraceptive methods are grouped together, and only the top three methods are displayed on the Sankey plot. Data are reported per patient. C, Sankey plot of changes in contraception methods by tier category from diagnosis to study inclusion. Data are reported per patient. Abbreviations: breast cancer (BC); intrauterine system (IUS); intrauterine device (IUD)

#### Changes in contraceptive methods

Between BC diagnosis and study inclusion, 381 patients (73.7%) had changed their contraception (Fig. 2B). Three main patterns of change were observed: (i) the number of women using hormonal contraception decreased strongly (from *n*=262 to *n*=15); (ii) the number of women using a copper IUD increased strongly (from *n*=41 to *n*=205); (iii) a substantial proportion of women with hormonal contraception at BC diagnosis had no contraception at study inclusion (85 of 262 (32.4%)). Most of the 268 patients with contraception both at diagnosis and at inclusion (84.7%) had continued to use the same method (*n*=122, 45.5%) or had switched to a higher tier of contraception (*n*= 105, 39.2%), whereas only 15.3% (*n*=41) had switched to a lower-tier method (Fig. 2C).

### Comparison of the prevalence of contraception and contraceptive methods between cases and controls

#### Population matching

For comparisons of the prevalence of contraception and contraceptive methods in BC survivors and controls, we matched the 517 BC patients with a control population of 3834 cancer-free volunteers included in the study on the basis of matching for age and parity in a 1:2 ratio, resulting in an overall population of 1551 women (cases *n*=517, controls *n* = 1034). After exclusion of the patients who were pregnant (*n*=103) or trying to conceive (*n*=170), 1278 women remained in cohort 2 for the analyses (case *n*=431, controls *n* = 847) (Fig. S1). Median age at inclusion in this study was 37.1 years. The controls were significantly more obese or overweight, and were more likely to be current smokers and to have concomitant comorbidity than the cases (Table1).

#### Prevalence of contraception and contraceptive methods

Overall, the prevalence of contraception did not differ between cases (340/431, 78.9%) and controls (666/847, 78.6%, *p*=0.97). Within the population of BC patients at risk of unintentional pregnancy, the factors associated with the use of contraception were younger age, the information about chemotherapy-induced ovary damage received at BC diagnosis, the information about contraception received at BC diagnosis, and anti-*HER2* treatment (TableS1). In the multivariate analysis, only information about chemotherapy-induced ovary damage (OR= 2.47 95% CI [1.39 - 4.37] and anti-*HER2* treatment (OR=2.46, 95% CI [1.14 - 6.16]) were significantly associated with the use of contraception.

The type of contraceptive method used differed significantly between cases and controls (Fig. 3A and Table2), with copper IUDs the major contraceptive method in cases, but with a lower frequency of use in controls (59.5% *versus* 25.0%, *p*<0.001). Contraceptive methods also differed significantly between cases and controls in terms of efficacy according to the tier classification (Fig. 3B) (*p*<0.001), and the use of hormonal versus non-hormonal methods (*p*<0.001) (Fig3C).

**Figure 3:**
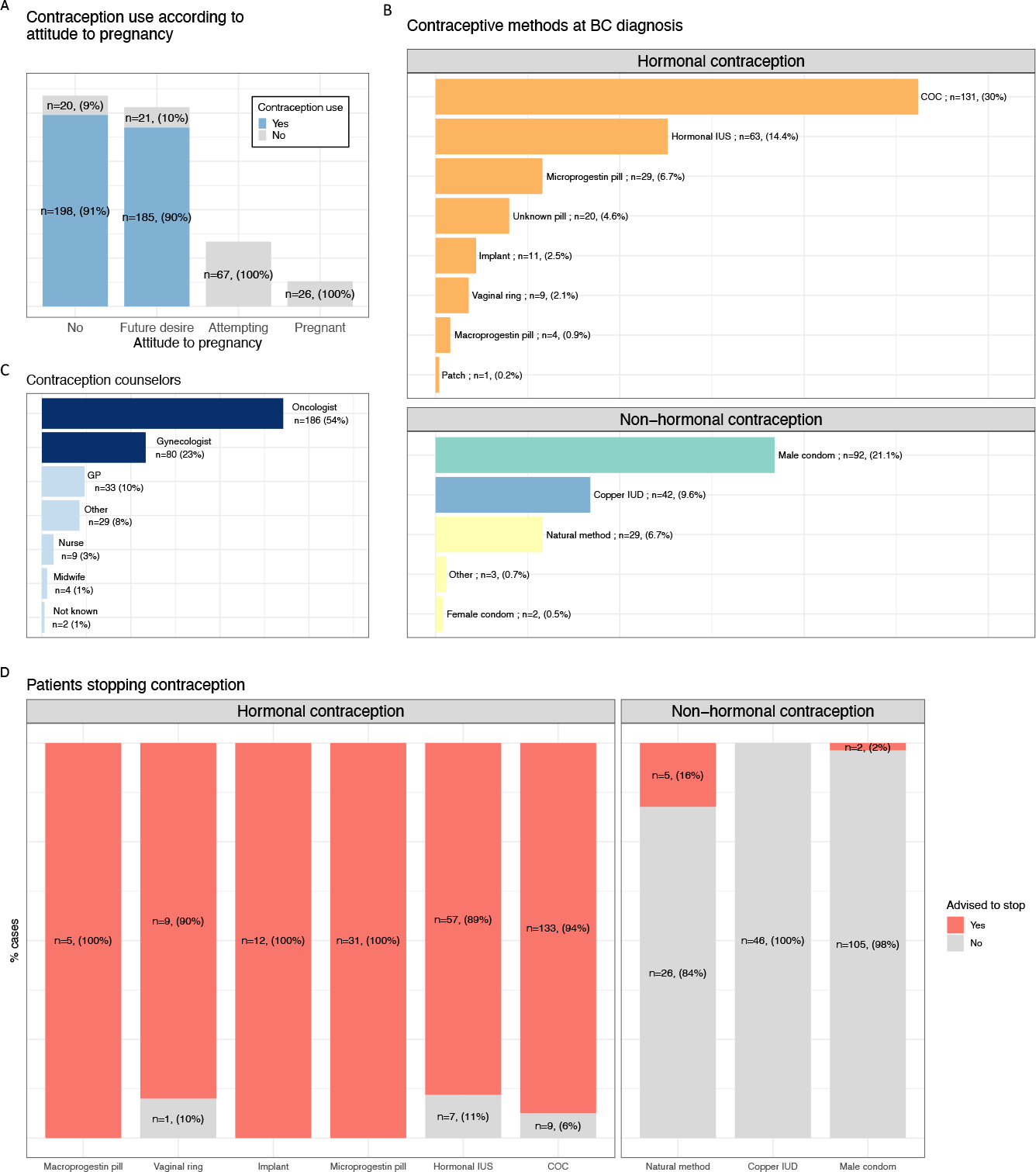
Comparison of contraceptive methods between cases and controls in the population of patients at risk of unintentional pregnancy. A, Contraceptive methods in cases and controls at inclusion in the study. B, Type of contraception, by tier category, at inclusion, for the cases and controls. C, Use of hormonal contraception by cases and controls at inclusion in the study. All data are reported per contraceptive method (one patient can use several methods). Abbreviations: combined oral contraceptive (COC); intrauterine system (IUS); intrauterine device (IUD)

### Definitive contraception

The prevalence of definitive contraception was low and did not differ significantly between cases and controls (21/358 (5.9%) *versus* 30/716 (4.2%) respectively, *p*=0.29) (Fig. S2A).

The vast majority of the definitive contraception methods used were female rather than male methods (66.7% *versus* 33.3%), with this tendency more marked among the cases than the controls (male methods: 19% for cases *versus* 44% for controls, *p*=0.17), although this difference was not statistically significant (Fig. S2B).

### Emergency contraception

The patients’ knowledge and use of emergency contraception are summarized in TableS2. All but 13 patients (1.0%) from this population were aware of the existence of emergency contraception. The proportion of women who had used emergency contraception was smaller for cases than for controls (42.0% *versus* 51.5%). In total, 617 women (48.3%) had used emergency contraception at least once in their lifetime (cases *n*=181 42%, controls *n*=437 51.3%), and 19 patients (4.4%) had used emergency contraception since BC diagnosis.

Only 10.3% of women were aware that an IUD could be used as an emergency contraceptive method. Overall, 38.1% of BC patients thought that oral emergency contraception was contraindicated due to their history of cancer

## Discussion

This large cohort study comparing BC survivors with age-matched controls provides important new insight into the contraceptive practices of BC patients.

Two thirds of the women received specific counseling at the time of BC diagnosis, mostly from medical oncologists. Several studies have shown that women with cancer lack knowledge about their contraceptive needs(Karaöz et al., 2010), or that healthcare practitioners do not address the reproductive needs of women with cancer (Patel et al., 2009; Peate et al., 2009). In the Fertility Information Research Study(Dominick et al., 2015), only half the survivors reported receiving contraception-related counseling, testing, prescriptions, or procedures after cancer diagnosis, despite a probably higher level of access to healthcare than for women without chronic medical conditions. Three quarters of the patients were asked to stop using their current mode of contraception. This is consistent with current WHO guidelines (WHO, 2015), according to which hormonal contraceptive use is considered to constitute a theoretically unacceptable health risk in patients with a history of BC (category 4). As a result, the proportion of women using hormonal contraception decreased strongly after BC diagnosis. Interestingly, this change in contraceptive use resulted in a switch to contraceptives of at least the same level of efficacy in most cases, with only a small subset of patients switching to a less effective contraceptive method.

We found that the overall prevalence of contraceptive use during follow-up was similar to that in matched controls from the general population. As summarized in Table 3, conflicting evidence has been published on this point. In an analysis of 295 cancers survivors from a US prospective cohort (Dominick et al., 2015), survivors were found to be less likely to use a contraceptive method than women in the general population (57% *versus* 69%, *p*=0.01), and less likely to use tier I–II contraceptive methods than the general population (34% *versus* 53%, respectively, *p*=0.01). In a study of 107 women of reproductive-aged diagnosed with cancer, Maslow *et al*. (Maslow et al., 2014) showed that these patients underused tier I/II contraceptive agents. Surprisingly, only four women in Maslow’s study reported using an IUD. Finally, in an American survey of 476 women under the age of 40 years and still menstruating after cancer treatment, Quinn (Quinn et al., 2014) found that sexually active cancer survivors had a three times higher risk of unintended pregnancy than women in the US general population. A history of previous BC (*n*=68 patients) was associated with a higher risk of unintended pregnancy (20.6%) than for any other group of respondents (10.5% (odds ratio 2.14, 95% CI 1.10–4.17 *p*=0.025). The discrepancies between published data and our findings may be explained by the origin of the patients, as the volunteers enrolled in the Seintinelles* network have a higher social status than allcomers, and social status is also strongly related to adequate contraception (Frippiat and Marquis, 2010). Finally, despite the higher prevalence of contraception in our study than in previous works, one in five of the women at risk of unintentional pregnancy declared no use of contraception, leaving room for improvement in the contraceptive coverage in this population.

The most frequently used contraceptive method after BC was, by far, the copper-IUD. Our study confirms that this method, which is recommended as the preferred option in guidelines (WHO, 2015), is feasible for BC survivors in real life. LNG-IUS were also used, but only in a very small subset of patients (*n*=9, 0.3%). The level of evidence concerning the risk of BC recurrence in patients using LNG-IUS remains very low (Fu and Zhuang, 2014; Gizzo et al., 2014; Trinh et al., 2008; Wong et al., 2013). However, LNG-IUs is still considered as contra-indicated after BC, also this last recommandation is not consensual (Vaz-Luis and Partridge, 2018).

Definitive contraceptive methods were used in no more than 5% of BC patients after treatment. This proportion is consistent with the rarity of definitive contraception in France (Vigoureux and Le Guen, 2018), possibly due to cultural barriers, a lack of knowledge, or such methods not being proposed by doctors. When definitive methods were chosen, they were predominantly of the female type, particularly in BC patients. Tubal ligation and Essure* are both considered to be more invasive than vasectomy, a minimally invasive technique that can be performed under local anesthesia. Definitive contraceptive methods are particularly appropriate for BC survivors not intending to have children in the future. Efforts should therefore be made to inform doctors; BC patients and their partners correctly, so that such contraceptive methods can be offered more widely. We also identified unfounded beliefs, such as the belief that hormonal emergency contraception is contraindicated in patients with a history of BC, which was held by up to one third of patients, highlighting the critical need for appropriate contraceptive counseling.

One of the strengths of this study is that it is the largest study to date providing data concerning contraception in BC cancer survivors, most previous studies having provided descriptive analyses in patients with cancers at various sites (Dominick et al., 2015; Hadnott et al., 2019; Maslow et al., 2014; Quinn et al., 2014). Women with BC are more likely to be older, to have already had several pregnancies and to be living with a partner or married, and educated than other young adults and teenage cancer survivors (Maslow et al., 2014). Together with the contraindication of hormonal contraceptive use, dedicated analyses in this specific population are of interest. The limitations of this study include the recruitment of women via online networks, which may have led to an overestimation of the prevalence of contraception, as most of the women in the FEERIC study came from high-level socioprofessional backgrounds.

This study calls for a number of actions. Medical oncologists were the first-line healthcare providers most frequently responsible for providing contraceptive counseling. As such, they should be offered specific training. Dedicated contraceptive counseling consultations onsite is also an appealing option, as the integration of such counseling has already proved effective for reproductive issues in fertility preservation (Peavey et al., 2017), and in the context of complex chronic diseases, such as cystic fibrosis in women (Rousset-Jablonski et al., 2020) and cardiovascular disease requiring anticoagulation (Bernard et al., 2018). Finally, sexual and reproductive health education programs (Aleksandra Korzeniewska-Eksterowicz et al., 2013), and survivorship care tools for improving reproductive health issues, including contraception (SCP-R, NCT02667626), could help to facilitate access to contraception and to ensure that patients are offered a wider range of contraceptive methods, including definitive methods, for which take-up remains poor.

## Data Availability

All data produced in the present work are contained in the manuscript

## Acknowledgments

The FEERIC study was funded by *Institut du Cancer* InCA, InCA-SHS, grant No. 2016-124, and is part of the *Young Breast Cancer Project*, funded by Monoprix*. The funder was not involved in study design, or in the collection, analysis, and interpretation of data, the writing of this article or the decision to submit it for publication. The authors thank all the study participants from the Seintinelles* Network and Lili Sohn who is the sponsor of the study.

## Legends figures and tables

### Figures and abbreviations

**Supplemental Figure 1:**
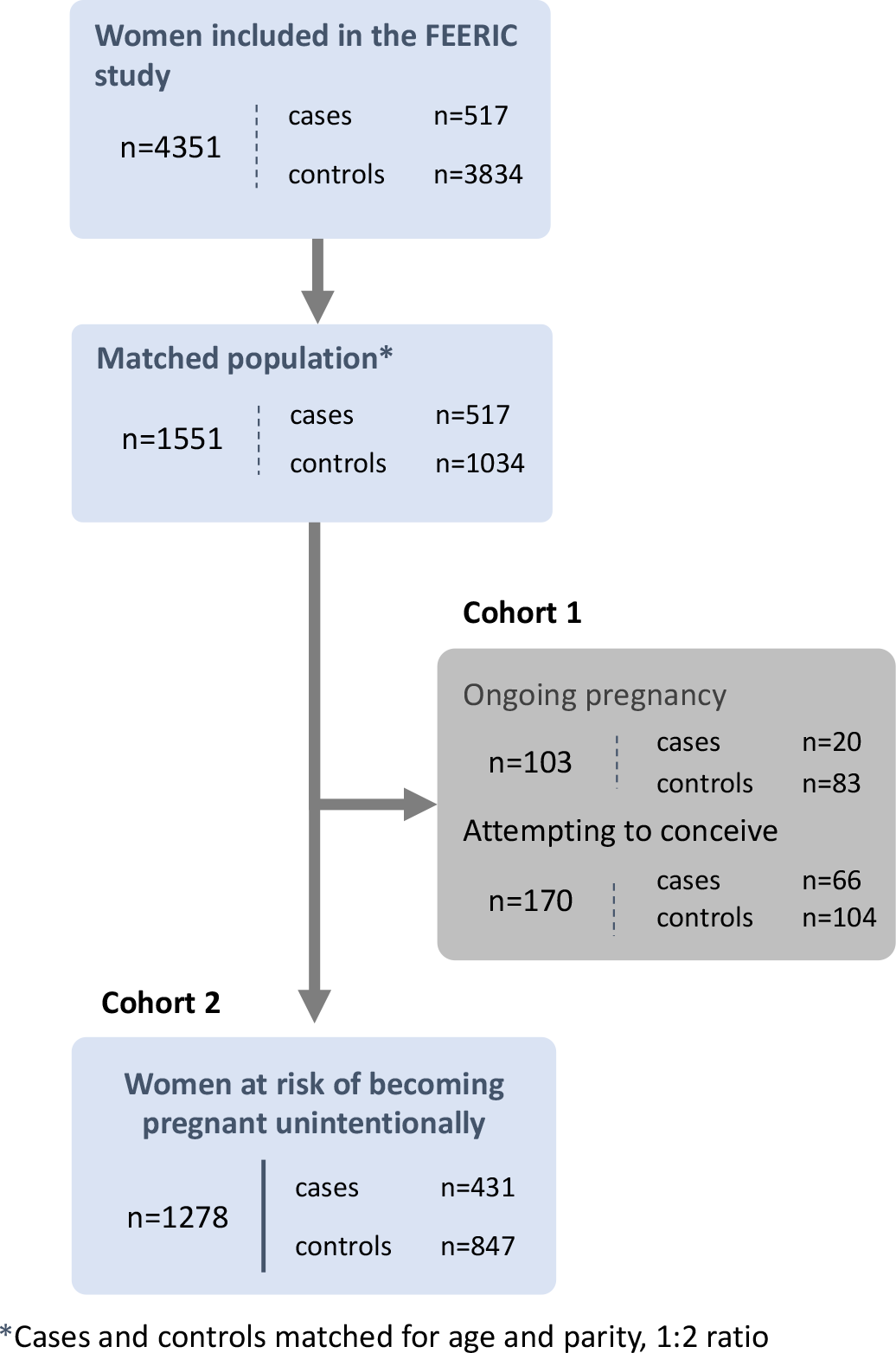
Flow chart for the study cohort

**Supplemental Figure 2:**
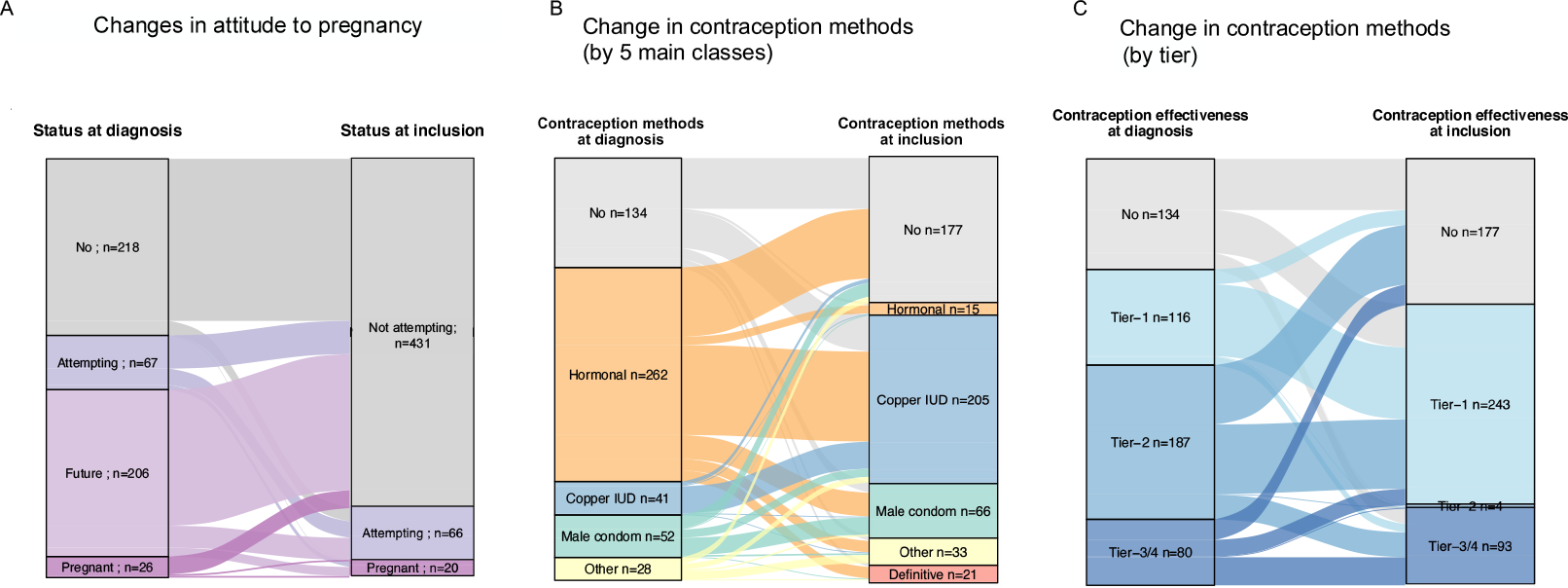
Use of definitive contraception in cases and controls A, Comparison of the use of definitive contraceptive methods between cases and controls. Data are reported per contraceptive method (one patient can use several methods). B, Comparison of the type of definitive contraceptive methods by cases and controls. Data are reported per contraceptive method (one patient can use several methods).

### Table titles and abbreviations

<u>Table 1: Characteristics of the women (cases and controls) in the population at risk of unintentional pregnancy.</u>

Each BC patient (case) was matched for age and parity to two volunteers (controls). We excluded 103 women currently pregnant and 170 women attempting to conceive from the analyses.

Abbreviations: breast cancer (BC); body mass index (BMI); socioprofessional category (SPC) “*n*” denotes the number of patients. Categorical variables are expressed as absolute numbers (percentages in brackets). Continuous variables are expressed as mean values, with the standard deviation in brackets. There were no missing data.

<u>Table 2: Methods of contraception used at study inclusion</u>

All data are reported per contraceptive method (one patient can use several methods). Abbreviations: intrauterine device (IUD); intrauterine system (IUS)

“*n*” denotes the number of patients. Categorical variables are expressed as absolute numbers, with percentages in brackets. Continuous variables are expressed as the mean value, with the standard deviation between brackets. There were no missing data.

<u>Table 3: Table summarizing studies analyzing contraception in BC survivors</u> Abbreviations:

<u>Supplemental Table 1: Factors associated with the use of contraception in patients with contraceptive needs</u> Abbreviations: socioprofessional category (SPC); breast cancer (BC); body mass index (BMI) “*n*” denotes the number of patients. Categorical variables are expressed as absolute numbers, with percentages in brackets. Continuous variables are expressed as the mean value, with the standard deviation in brackets.

Analysis performed for cases only, because too many variables of interest were missing for multivariate analysis on the whole population. There were no missing data.

<u>Supplemental Table 2: Knowledge and use of emergency contraception in cases and controls</u> Abbreviations: intrauterine device (IUD); breast cancer (BC)

“*n*” denotes the number of patients. Categorical variables are expressed as absolute numbers, with percentages in brackets. Continuous variables are expressed as the mean value, with the standard deviation in brackets. For non-normal continuous variables, the median value is reported, with the interquartile range in brackets.

Missing data: type of emergency contraception pill, *n*=2; type of emergency contraception pill used since BC diagnosis, *n*=3.

## Notes

### Competing Interest Statement

The authors have declared no competing interest.

### Author Declarations

Data were collected via self-administered online questionnaires released through the Seintinelles* research platform. Seintinelles* is a collaborative social network created in 2012 to accelerate the recruitment of French volunteers for cancer research studies, by connecting researchers with men and women of various ages, social and medical backgrounds, with or without a history of BC, willing to participate in research studies. The scientific board of the Seintinelles approved the FEERIC project in December 2015, and the ethics board of Sud Ouest Outre Mer II approved the project on October 5, 2017.

